# Rapid transmission of coronavirus disease 2019 within a religious sect in South Korea: a mathematical modeling study

**DOI:** 10.1101/2021.08.05.21261683

**Authors:** Jong-Hoon Kim, Hyojung Lee, Yong Sul Won, Woo-Sik Son, Justin Im

## Abstract

Rapid transmission of coronavirus disease 2019 (COVID-19) was observed in the Shincheonji Church of Jesus, a religious sect in South Korea. The index case was confirmed on February 18, 2020 in Daegu City, and within two weeks, 3,081 connected cases were identified. Doubling times during these initial stages (i.e., February 18 – March 2) of the outbreak were less than 2 days. A stochastic model fitted to the time series of confirmed cases suggests that the basic reproduction number (*R*_0_) of COVID-19 was 8.5 [95% credible interval (CrI): 6.3, 10.9] among the church members, whereas (*R*_0_ = 1.9 [95% CrI: 0.4, 4.4]) in the rest of the population of Daegu City. The model also suggests that there were already 4 [95% CrI: 2, 11] undetected cases of COVID-19 on February 7 when the index case reportedly presented symptoms. The Shincheonji Church cluster is likely to be emblematic of other outbreak-prone populations where *R*_0_ of COVID-19 is higher. Understanding and subsequently limiting the risk of transmission in such high-risk places is key to effective control.

**Highlights:**

- Basic reproduction number (*R*_0_) of COVID-19 in a religious community of Shincheonji Church of Jesus was estimated to be 8.5 [95% credible interval (CrI): 6.3, 10.9], which is more than 4 times larger than the general population (*R*_0_ = 1.9 [95% CrI: 0.4, 4.4])
- There were estimated 4 [95% CrI: 2, 11] undetected cases when the index case from the religious community reported symptom on February 7.
- The Shincheonji Church cluster is likely to be emblematic of other outbreak-prone populations where *R*_0_ of COVID-19 is higher. Understanding and subsequently limiting the risk of transmission in such high-risk places is key to effective control.

## 1. Introduction

Coronavirus disease 2019 (COVID-19) has become a global pandemic since it was first reported in Wuhan, China in December 2019 with the name of novel coronavirus disease (Li et al., 2020a). The causative agent, severe acute respiratory syndrome coronavirus 2 (SARS-CoV-2), transmits mainly through human-to-human contact (Chan et al., 2020), which can happen even during the infector is asymptomatic (Rothe et al., 2020; Yu et al., 2020). Infection with the virus causes diseases with varying degree of symptoms including death (Fu et al., 2020; Guan et al., 2020). Infection mortality ratio is lowest among children aged between 5 and 9 years and increases loglinearly with age (O’Driscoll et al., 2021).

One key characteristic of COVID-19 pandemic is that transmission events in high-risk settings such as super-spreading events (SSEs) contribute to most transmissions (Adam et al., 2020; Lemieux et al., 2020). The risk of COVID-19 transmission is believed to high in places with high occupancy and poor ventilation (Jones et al., 2020). One extreme example is the outbreak in the Diamond Princess cruise ship, where 17% (619/3711) of the passengers were infected from January 25 to February 20, 2020 (Russell et al., 2020). Other examples include transmission events in bars and wedding (Adam et al., 2020) in Hong Kong, nursing homes in U.S. (Chen et al., 2021), telemarketers working in group in closed places (Park et al., 2020) and fitness classes (Jang et al., 2020) in South Korea, and also religious gatherings, which we describe below.

Explosive spread of COVID-19 was observed in the Shincheonji Church of Jesus (“Shincheonji”), a religious sect in South Korea. The index case was confirmed on February 18, 2020 and within two weeks, 3,081 connected cases were identified (Korea Disease Control and Prevention Agency, 2020a). A simple calculation reveals that the outbreak size doubled in less than every 2 days (14/log_2_(3081) ≈ 1.21), which is smaller than doubling times reported in the early stages of the COVID-19 outbreak in China (2.5 and 3.1 days in Hubei Province and Hunan Province, respectively) (Muniz-Rodriguez et al., 2020b), Spain (2.8 days) (Guirao, 2020), the US (2.7 days) (Lurie et al., 2020), and Korea (2.8 – 10.2 days) (Shim et al., 2020b). A total of 6,684 confirmed cases were reported in Daegu City as of March 31, 2020 of which 4,467 (66.8%) were Shincheonji members, representing close to half (47.9%, 4,467/9,334) of the city’s total Shincheonji membership.

Previous studies highlighted that COVID-19 transmissions involve SSEs (Xu et al., 2020), which can play a key role in sustained community transmissions (Adam et al., 2020; Lemieux et al., 2020). However, there have been no attempts to model the dynamics of COVID-19 transmission within high-risk settings and their interaction with the general community. In this study, we modeled the outbreak in the Shincheonji community while accounting for its interaction with the rest of population. We used the stochastic model to account for the stochastic nature of the transmission events. We used the model to explore the differences in the basic reproduction number (*R*_0_) between the high-risk setting and the general community, and quantify uncertainties related to the initial conditions and dynamics of transmission under the dynamic intervention programs.

## 2. Materials and Methods

### 2.1 Backgound on the Shincheonji Church of Jesus

Shincheonji was founded by Man-hee Lee in 1984 and has approximately 245,000 members including 30,000 foreigners (Chung and Hill, 2020). At Shincheonji gatherings, worshipers used to sit close together on the floor and facial coverings, such as glasses and face masks, are forbidden. Members were expected to attend services despite illness (Choe, 2020). The index case of the Daegu City outbreak was identified as a Shincheonji member and some 1,000 people were reported to have attended worship together (Yonhap, 2020). Further tracing of church members identified clustering in apartment complexes. Of 142 residents in a particular Daegu apartment block, 94 (66%) were Shincheonji members of whom 46 (38.9%) tested positive for the virus (Myung, 2020).

### 2.2 Data

Time series of patients confirmed with COVID-19 in Shincheonji community and the overall Daegu City over the period of 19 February – 31 March 2020 was compiled based on the daily reports from Korea Disease Control and Prevention Agency (KDCA) (Korea Disease Control and Prevention Agency, 2020a) (Figure 1). The reports provide the number of cases confirmed with SARS-CoV 2 based on reverse transcription polymerase chain reaction (RT-PCR) by category (Shincheonji or non-Shincheonji). We made some adjustments to the existing data before we fit the model. First, in the beginning of the outbreak, KDCA provided both daily and cumulative numbers of cases confirmed for SARS-CoV 2, which did not agree always. If there is a discrepancy between these numbers, we prioritized cumulative numbers as this figure was reported continuously throughout the outbreak. Second, data were missing for some days for the number cases for Shincheonji members. We imputed missing values using the cubic spline method (Figure S1 in the Supplementary Material).

### 2.3. Doubling time

The epidemic doubling time (*T*_*d*_) represents the duration in which the cumulative incidence doubles. Assuming exponential growth with a constant epidemic growth rate (*r*), the epidemic doubling time can be calculated by the following equation (Anderson et al., 2020; Lurie et al., 2020; Muniz-Rodriguez et al., 2020a)

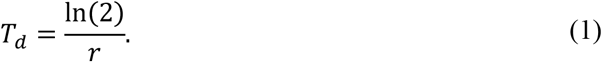

Epidemic growth rate (*r*) may be estimated based on the data. For example, *r*(*t*) can be estimated by the following equation:

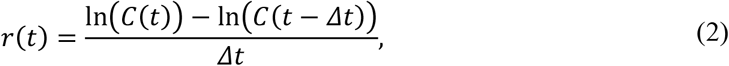

where *C*(*t*) indicates the cumulative number of infected people at time *t* and *Δt* is the duration over which *r*(*t*) is assumed to be constant. *r*(*t*) can be calculated over the fixed time interval (e.g., 1 day or 1 week) (Ebell and Bagwell-Adams, 2020; Patel and Patel, 2020) or variable time intervals (e.g., days on which the number of cases doubles, quadruples, etc.) (Muniz-Rodriguez et al., 2020a; Shim et al., 2020b). We calculated doubling based on prior 7 days or 1 day from 18 February to 5 March 2020, when the epidemic peaked and no further doubling of cumulative number of cases occurred onward.

The basic reproduction number (*R*_0_) is defined as the average number of secondary cases caused by a single infected case in an entirely susceptible population and it provides sufficient information to produce doubling times in the beginning of an outbreak. However, estimating *R*_0_ requires additional information such as generation time or developing a mechanistic model, and its estimates come with higher degree of uncertainty (Anderson et al., 2020). Calculating doubling times requires fewer assumptions and also allows us to compare our results with estimates from different settings where doubling times, but not reproduction numbers, are available.

### 2.4. Mechanistic model of COVID-19 transmission

We developed a stochastic model of COVID-19 transmission within the Shincheonji community and the overall population of Daegu City. The model includes six disease states: susceptible (*S*), exposed but not infectious (*E*), pre-symptomatic but infectious (*P*), symptomatic and infectious (*I*), asymptomatic but infectious (*A*), confirmed and isolated (*C*), and recovered (*R*). The model includes two patches to model Shincheonji and non-Shincheonji people, separately. Transmission rates may differ for each patch and person from one patch may infect people from the other patch. (Figure 2).

This modeling framework of mixing between two distinct sub-populations has been adopted in previous works, ranging from sexually transmitted diseases (Koopman et al., 1988) to vector-borne diseases such as dengue (Lee and Castillo-Chavez, 2015), where formulations for mixing between patches vary. We adopted the formulation used in the work on modeling transmission of cholera between hotspot and non-hotspot areas (Azman and Lessler, 2015). Mixing between two sub-populations are defined by the 2 × 2 contact matrix,

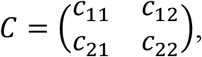

where *c*_*ij*_ indicates the fraction of time that individuals from patch *i* spends in patch *j*. Next, we impose two conditions on the matrix *C*:

i. Individuals must reside in either of the two patches, i.e., *c*_*i*1_ + *c*_*i*2_ = 1 for *i* =1 (Shincheonji) and 2 (non-Shincheonji).
ii. The population in each patch remains constant, i.e., *c*_12_*N*_1_ = *c*_21_*N*_2_, where *N*_1_ and *N*_2_ represent population size for patch 1 and 2, respectively.

The above conditions may transform the contact matrix *C* to the following form:

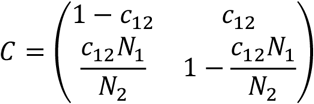

containing only one unknown parameter *c*_12_.

The force of infection for individuals from patch *i* at time *t, λ*_*i*_(*t*), is defined as follows:

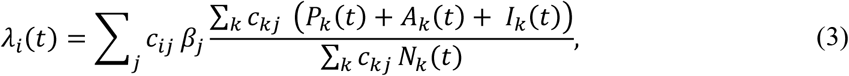

where *β*_*j*_ indicates local transmission rate in patch *j* and *I*_*k*_(*t*) indicates number of infectious individuals from patch *k*.

The transitions between states are modeled using an explicit tau-leap algorithm (Gillespie, 2001) to account for stochasticity of the infection transmission process. The number of susceptible people in patch *i* at time *t* + Δ*t, S*_*i*_(*t* + Δ*t*), is written as follows:

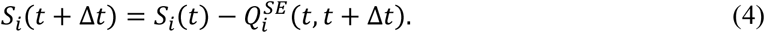

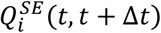 represents the number of people who transit from state *S* to state *E* from *t* to *t* + Δ*t* in patch *i* and is a random variable with binomial distribution:

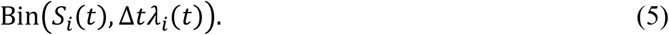

That is, it is represented as an integer varying between 0 and *S*_*i*_(*t*). For states from which more than one potential transition exist (e.g., *P* to either *A* or *I*), multinomial distributions were applied. For instance, the number of people transit from *P* to either *I* or *A* are given as follows:

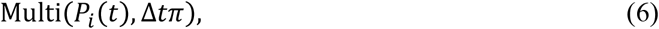

where *π* is a vector given as

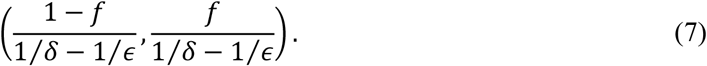

The first element of *π* indicates a probability of transition from *P* to *I* and the second element indicates the probability of transition from *P* to *A*. The number of people in other states (i.e., *E, A, I, C, R*) at time t can be described similarly. The model was implemented in a combination of R and C++ languages, in which the core transmission model part is expensive and was written in C++. All the computer codes that generate the results in this paper are available at the author’s GitHub repository (Kim, 2021).

### 2.5. Modeling intervention program

To account for intensification of the intervention such as case isolation and contact tracing with subsequent testing during the outbreak, we assumed case isolation rate (1 / mean time between symptom onset and case isolation) and transmission rate of the infectious people per unit time change over time. Specifically, we assumed that the case isolation rate, α(*t*), starts increasing on February 20 from the initial value of *α*^init^ when 4,474 out of 9,334 Shincheonji members were identified and were asked to self-isolate. During model fitting, we let data suggest the duration of intervention, *d* in day, which is the time required for the case isolation rate to reach its minimum, *α*(*t*) = *α*^final^ for *t* > February 20 + *d*. We assumed that the mean time between symptom onset and case isolation linearly decreases over the intervention period *d*. In other words, α(*t*) is formulated as follows:

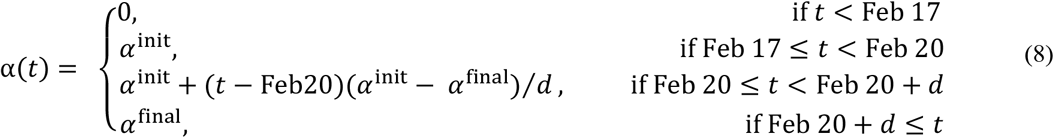

where *α*^final^ is assumed to be 1 day based on the experiences in Busan City in Korea and α(*t*) is assumed to be zero before February 17 when the index case was detected.

Similarly, transmission rate per unit time at time *t, β*_*i*_(*t*) for *i* = 1 (Shincheonji members), 2 (non-Shincheonji people in Daegu City), is assumed to linearly decrease during the intervention period.

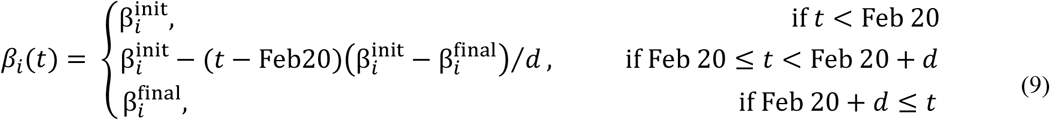

Here, 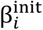 and 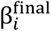 indicate the transmission rate per unit time before the intervention and after the intervention measures fully take effect, respectively. They can be derived once *R*_0,*i*_ and *R*^final^ are given as:

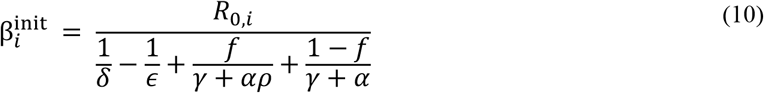

and

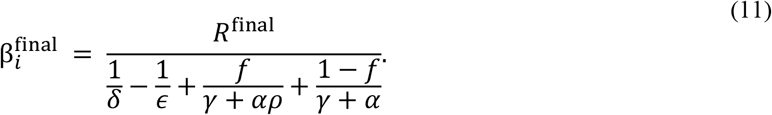

### 2.6. Parameter estimation

Our model of COVID-19 transmission requires 15 parameters (Table 1). We divided the model parameters into three classes depending on our belief on their relative certainty. The first class includes parameters related to the natural history of infection and population size and we deemed that available parameter estimates are reliable. For these parameters, we used their point estimates based on analyses of data on COVID-19 transmissions in Korea or China. For the second class, which includes parameters related to intervention programs, we used our best guesses based on supporting evidence but still acknowledged their uncertainty. Therefore, we analyzed the models under various assumptions on their values within some pre-specified ranges. Finally, we defined six parameters that are critical for characterizing dynamics of COVID-19 transmission in Shincheonji members and non-Shincheonji people. We estimated these parameters by fitting the model to daily confirmed COVID-19 cases of Shincheonji members and non-Shincheonji people.

Estimation of parameters θ = (*R*_0,1_, *R*_0,2_, *I*_0_, *c*_12_, *d, R*^final^) was based on Approximate Bayesian Computation Sequential Monte Carlo (ABC-SMC) (Minter and Retkute, 2019). The ABC is a method for approximating posterior distributions given data *D, p*(θ|*D*), by accepting proposed parameter values when the difference between simulated data *D*^*^ and *D, d*(*D, D*^*^), is smaller than tolerance ϵ:

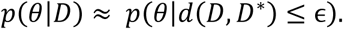

For our model, *d*(*D, D*^*^) is defined as the sum of the squared differences in daily confirmed cases over the outbreak of duration *T* days, that is,

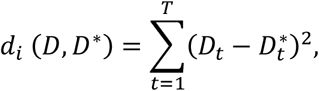

for Shincheonji (*i* = 1) and non-Shincheonji (*i* = 2). Here, *D*_*t*_ and 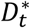 represent observed daily confirmed cases and model predicted values at time day *t*, respectively. ABC-SMC was designed to increase efficiency of the ABC method and ABC is applied in a sequential manner by constructing intermediate distributions, which converge to the posterior distribution. Tolerance ϵ is gradually decreased and each intermediate distribution is obtained as a sample that is drawn with weights from the previous distribution and then perturbed through a kernel *K*(θ|θ^*^). The kernel helps keep the algorithm from being stuck in local optimum while maintaining the efficiency of the ABC-SMC method. Minimally informative uniform distributions were used as prior distributions and estimation procedure was repeated for ten different random seeds. The resulting distribution was summarized as median, 50% credible intervals (CrI; interval between 25% and 75% percentiles) and 95% CrI (interval between 2.5% and 97.5% percentiles). More details of the algorithm such as prior distribution for each parameter, the number of steps, the tolerance values for each step, perturbation kernel appear in the Supplementary Material.

## 3. Results

### 3.1 Doubling time

Over the period of February 18 – March 5, during which doubling of confirmed cases occurred 12 times, doubling times were <1 day in the beginning and increased subsequently with daily doubling time presenting higher variability for both Shincheonji and non-Shincheonji values (Table 2). Doubling times calculated over sliding one-week intervals remained shorter than 3 days for the most part for both Sincheonji and non-Shincheonji population.

### 3.2 Comparison between observations and the mechanistic model

Our fitted model projects the trajectory of number of daily and cumulative confirmed cases in Shincheonji and in the rest of the population of Daegu City (Figure 3(a)-(d)). The model correctly projects the decreasing trends in both patches after reaching the peak on around March 3, 2020. However, for the non-Shincheonji, daily new cases are underestimated toward the end of the outbreak.

*R*_0_ were estimated to be quite different across two patches (Figure 3(e)). The local reproduction number in the patch representing Shincheonji members, *R*_0,1_, was estimated to be 8.54 [95% credible interval (CrI): 6.30, 10.95] whereas the local reproduction number in the patch representing non-Shincheonji members, *R*_0,2_, was estimated to be 1.87 [95% CrI: 0.38, 4.40]. The time taken for the intervention program to have exerted highest effect, *d*, is around 9.02 days [95% CrI: 7.85, 10.45], which leads to both reduced transmission rate per unit time and reproduction numbers (*R*^final^ = 0.34 [95% CrI: 0.18, 0.53]). The model also suggests that there were infectious people already when the first cases was symptomatic on February 7 (*I*_0_= 4 [95% CrI: 2, 11]). The proportion of time that a person from Shincheonji members spends mixing with non-Shincheonji people, *c*_12_, was estimate be around 0.14 [95% CrI: 0.05, 0.22]. Posterior distribution of parameters based on 2,000 samples obtained from 10 different random seeds and two-way correlations appear in Supplementary Material (Figure S2).

## 4. Discussion

Rapid transmission of COVID-19 within the Shincheonji community is likely to have been facilitated by high intensity contact between individuals gathering during services and in residential areas. Our mathematical modeling analyses quantify the rapid spread of COVID-19 in Daegu City driven by a community of Shincheonji members. The median *R*_0_ among Shincheonji members (*R*_0,1_) was 8.5, which is over 4-fold higher than what was estimated for the rest of the population in Daegu City (*R*_0,2_ =1.9). While the *R*_0_ in the Shincheonji community is higher than estimates from most transmission hotspots (e.g., in China (Alimohamadi et al., 2020; Imai et al., 2020; Riou and Althaus, 2020; Wu et al., 2020; Zhao et al., 2020b) and Korea (Bae et al., 2020; Choi and Ki, 2020; Ki, 2020; Shim et al., 2020a)), such high *R*_0_is not unusual in particular considering that *R*_0_ can be different depending on the local settings with varying contact rates (Temime et al., 2020). Studies do report that *R*_0_ estimates of COVID-19 that are comparable or even higher than our estimates for the Shincheonji community. During periods of intensive social contacts near the Chinese New Year in China, *R*_0_ was estimated to be 6 (Sanche et al., 2020; Tang et al., 2020). Also, *R*_0_ estimates were around 5 among those traveled from Wuhan and were subsequently confirmed in other countries (Zhao et al., 2020a), and around 7 during the initial growth phase in the UK (Dropkin, 2020). In an extreme setting such as the Diamond Princess ship, much higher estimates (*R*_0_ = 14.8) were reported (Rocklöv et al., 2020). Although the previous studies that included data on the outbreak in Shincheonji community report smaller *R*_0_ estimates (Choi and Ki, 2020; Shim et al., 2020a) than our estimates, the difference might stem from that prior studies did not model the Shincheonji community separately from the rest of the population and therefore measured the *R*_0_ averaged across sub-population that are highly heterogeneous.

Although estimated daily doubling times show some variability (e.g., 14 days on February 28 and 69.6 days on March 1 for the non-Shincheonji population), they are short overall, which indicates rapid growth of the outbreak, and are compatible with estimates from other settings. Daily doubling times were lower than one day in the beginning of the outbreak and this is similar to the estimates from several regions in China (Muniz-Rodriguez et al., 2020a). The study by Shim *et al*. (Shim et al., 2020b) used the dataset from Daegu City, Korea including Shincheonji population produced the doubling time of 2.8 days [95% CI: 2.5, 4.0]. Our daily doubling time estimates averaged over the period of February 18 – March 5 is 2.9 days and is consistent with the study. The period from February 18 to March 5 is likely to have been used in the study by Shim *et al*. because the authors calculated the doubling times on the days when the reported cases doubled and during the period of February 18 – March 5 the number of cases doubled 12 times and no further doubling occurred since then. The study by Lee *et al*. (Lee et al., 2020) used similar data, but reported seemingly inconsistent findings, doubling time of 2.9 days for the first week and 3.4 over the period around February 18 – March 4 considering that our estimate averaged over the first week is 0.9 day. One likely reason for this difference is that Lee *et al*. calculated the doubling time using the cumulative incidence estimated from a logistic model that used the initial value (i.e., number of infected people on February 18) as a free parameter. Figure 2D from their study indicates that the number of infected people on 18 February is much larger than 1 and this might have led to the higher doubling time than our estimates. This may also explain why Lee *et al*. estimates for a similar period (i.e., February 18 – March 5) is higher than our estimates and those by Shim *et al*. (Shim et al., 2020b).

The relationship between the doubling time and the *R*_0_ provides two insights on our inferences on *R*_0_. For an *SEIR* model, there exists an algebraic formula that describes the inverse relationship between initial epidemic growth rate and *R*_0_ (Ma, 2020; Ma et al., 2014). This inverse relationship suggests that short doubling times during the early phase of the epidemic we calculated using the growth rates are consistent with high *R*_0_ for Shincheonji we estimated using the stochastic dynamic transmission model (Table S2 in the Supplementary Material). On the other hand, while doubling times may be reduced and imply high *R*_0_ for non-Shincheonji people as well, such short doubling times can arise through mixing (i.e., positive *c*_12_) with Shincheonji of high *R*_0_ even if the *R*_0_ for the non-Shincheonji people are not as high.

Parameters around asymptomatic infections of COVID-19 are largely unknown (Fox et al., 2020) and we tested the sensitivity of our inferences to our assumptions on two parameters related to asymptomatic infection, namely the proportion of asymptomatic infection, *f*, and relative rate of isolation of asymptomatic people, *ρ* (Figure S3 in Supplementary Material). *R*_0_ for Shincheonji people, *R*_0,1_, and the final reproduction number, *R*^final^, showed a slight increase with increasing *f* or decreasing *ρ* while other parameter estimates remain relatively constant. We also tested the sensitivity of our parameter estimates to *α*^final^ , maximum rate of isolation near the end of the outbreak. *α*^final^ showed an inverse relationship with other intervention-related parameters such as duration of intervention *d*, and reproduction number at the end of the outbreak, *R*^final^. Overall, while there are some quantitative differences in our parameter estimates in response to the change in our assumptions on fixed parameters, *R*_0,1_ was always over 4-fold higher than *R*_0,2_.

While the first case was confirmed on February 18 for the Shincheonji outbreak, it was later revealed that the first case had symptoms on February 7 and even earlier transmission events were also suspected (Korea Disease Control and Prevention Agency, 2020b). This finding is consistent with our model analyses, which suggest there were 4 [95% CrI: 2, 11] infectious people on February 7. These undetected cases are likely to have contributed to the explosive outbreak in the Shincheonji community. Studies suggest that a substantial fraction of all SARS-CoV-2 infections were undetected. For Korea, it was suggested that the number of undetected cases may be larger than the number of detected cases (Lee et al., 2021). A study suggests that > 80% of all infections were undocumented during the initial spread in China (Li et al., 2020b). In France, over the period of 7 weeks since 28 June 2020 after the first lockdown, it was estimated that around 93% of all symptomatic cases were undetected initially and later around 69% of symptomatic cases were undetected by the time when case ascertainment improved (Pullano et al., 2021).

We have shown that SARS-CoV 2 has disproportionately affected a religious community generating a large cluster of linked cases in Korea. Similar large clusters of cases in high-risk settings have been observed in Korea and elsewhere. In Korea, many similar outbreaks in high-risk settings have been reported in the news including the outbreak in a dance class (Jang et al., 2020) and a call center (Park et al., 2020). In Singapore, a total of 247 cases were confirmed as of March 17, 2020 and six clusters including the spread in a hotel and in a church accounted for 45.3% of the total cases (Tariq et al., 2020). In Hong Kong. 1,038 cases were confirmed from January 23 to April 28, 2020 and among them, 51.3% of cases were associated with large clusters. Such social settings as bars, restaurants, weddings, and religious sites appeared at increased risk of large outbreaks (Adam et al., 2020).

One limitation of our analyses is that the model was fit to date of case confirmation because the date of symptom onset, which is more closely related with the date of infection, was not available. The daily number of confirmed cases can abruptly change depending on the intensity of intervention measures, of which the dynamics may not be consistent with disease transmission process. This means using the data on case confirmation under dynamics intervention measures is challenging. We tried to mitigate this difficulty by incorporating the dynamics of intervention programs by assuming that the start date and duration of enhanced case detection vary while the case detection rate increases over time and let the data suggest the values for those parameters.

## 5. Conclusions

The potential for large variations in *R*_0_ for COVID-19 has important implications for the design and effectiveness of control strategies. The efficacy of components of intervention programs, such as contact tracing and physical distancing, is dependent on various environmental and societal factors (e.g., large gatherings, physical proximity, high risk behaviors such as singing, etc.) that influence the transmissibility of disease. Our analyses provide important insights that in order to minimize the risk of sudden outbreaks, efforts to identify and preempt high transmission scenarios will be key to controlling the spread of the COVID-19. Understanding and subsequently limiting the risk of transmission in high-risk places such as the Shincheonji Church cluster in Korea is key to effective control of COVID-19 transmission.

## Data Availability

All of the data referred to in the manuscript is publicly available.

## Funding

This research was partly supported by Government-wide R&D Fund project for infectious disease research (GFID), Republic of Korea (grant number: HG18C0088) and National Institute for Mathematical Sciences (NIMS) grant funded by the Korean Government (NIMS-B21910000).

## Acknowledgments

All authors acknowledge discussions with the members of the Research and Development on Integrated Surveillance System Development for Early Warning of Infectious Diseases of Korea.

## Supplementary Material

### Data source

Figure S1 shows the cumulative cases before and after imputation. We used the cubic spline method provided in the imputeTS package of R.

**Figure S1.**
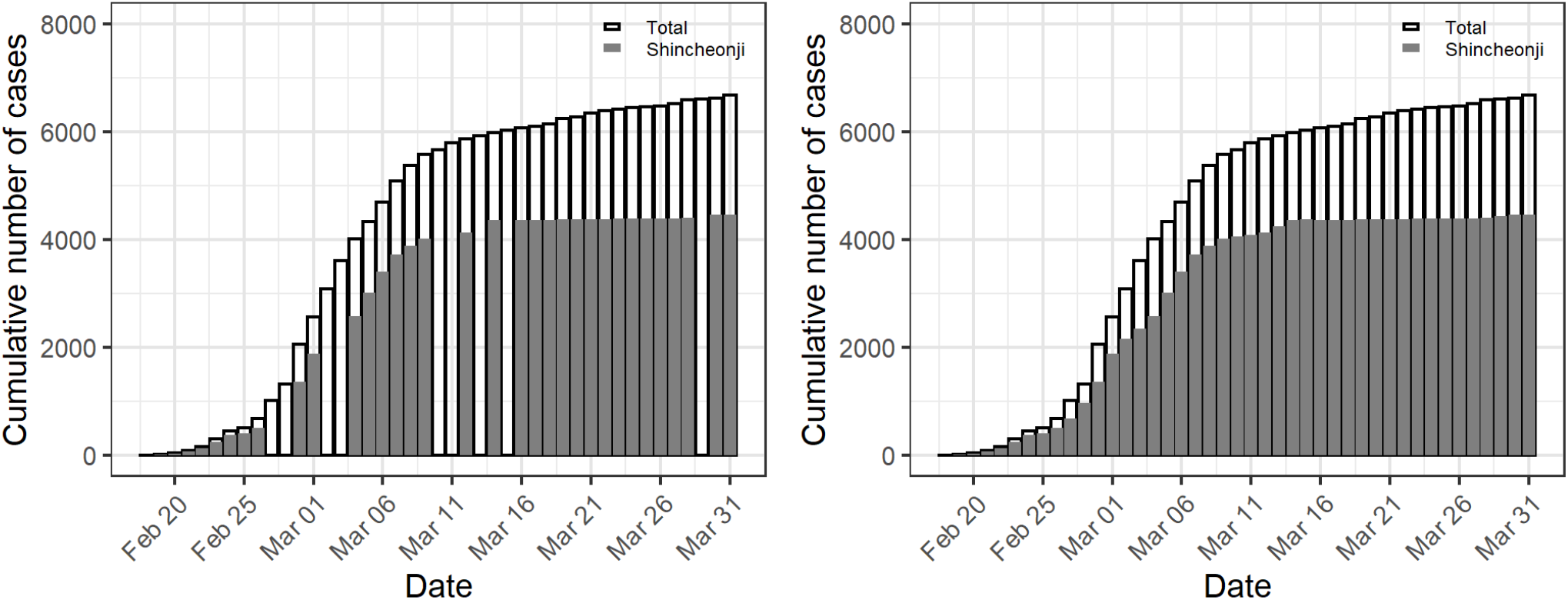
Cumulative number of cases in Shincheonji community and the overall Daegu before (left panel) and after imputation (right panel).

### Model equations

The model is implemented using a tau-leap algorithm. The number of people who transit from state *x* to state *y* over the time interval from *t* to *t* +Δ*t* in patch 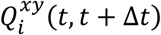 , is defined as follows:

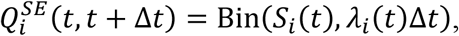

where Bin(*n, p*) represents binomial and with parameters *n* and *p*, respectively.

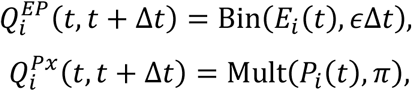

where Multi(*n*, π ={π_1_, π_2_}) indicates multinomial distribution with parameters *n* and π. π is given as follows:

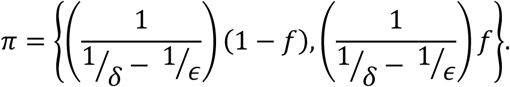

The first element of *π* indicates a probability for *x* = *I* (i.e., transition from *P* to *I*) and the second element indicates the probability of transition from *P* to *A* (*x* = *A*).

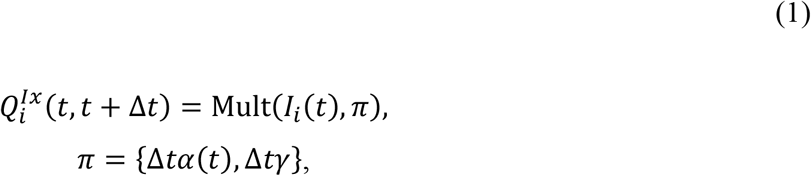

The first element of *π* indicates a probability for *x* = C (i.e., transition from *I* to *C*) and the second element indicates the probability of transition from *I* to *R* (*x* = *R*).

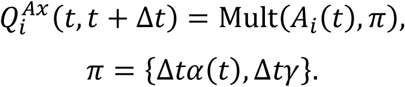

The first element of *π* indicates a probability for *x* = C (i.e., transition from *A* to *C*) and the second element indicates the probability of transition from *A* to *R* (*x* = *R*).

The number of people in each state at time *t* + Δ*t* can be described using the terms defined above:

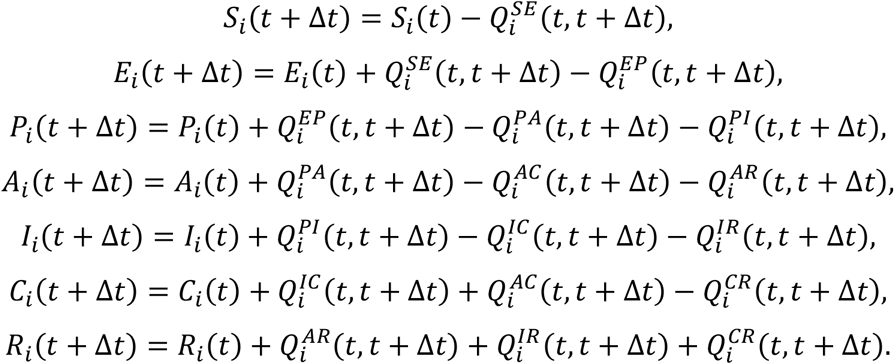

The model comprises two sets of above equations that describe two patches (i.e., a community of Shincheonji members and the non-Shincheonji people in Daegu City) and these equations are linked through the force of infection function, *λ*(*t*), which is defined in the main text.

**Table S1.** Delay from onset of symptoms to isolation during the COVID-19 outbreak in Busan City, Korea.

| Date | Median | Mean | Number of cases |
| --- | --- | --- | --- |
| 2/6/2020 | 17.0 | 17.0 | 1 |
| 2/16/2020 | 8.0 | 8.0 | 1 |
| 2/17/2020 | 5.5 | 5.5 | 2 |
| 2/19/2020 | 4.0 | 3.7 | 7 |
| 2/20/2020 | 2.5 | 2.6 | 8 |
| 2/21/2020 | 1.5 | 2.2 | 6 |
| 2/22/2020 | 1.0 | 2.0 | 9 |
| 2/23/2020 | 2.0 | 1.8 | 4 |
| 2/24/2020 | 0.0 | 0.7 | 3 |
| 2/25/2020 | 1.5 | 1.5 | 2 |
| 2/26/2020 | 1.5 | 1.8 | 4 |
| 2/27/2020 | 1.0 | 2.0 | 3 |
| 2/28/2020 | 3.0 | 2.3 | 3 |
| 2/29/2020 | 6.0 | 6.0 | 2 |
| 3/1/2020 | 2.0 | 2.0 | 1 |
| 3/2/2020 | 3.0 | 3.0 | 1 |
| 3/4/2020 | 3.0 | 3.0 | 1 |
| 3/6/2020 | 8.0 | 8.0 | 1 |
| 3/9/2020 | 3.0 | 3.0 | 2 |
| 3/12/2020 | 1.0 | 1.0 | 1 |
| 4/8/2020 | 10.0 | 10.0 | 1 |
| 5/6/2020 | 3.5 | 3.5 | 2 |
| 5/10/2020 | 2.0 | 2.0 | 1 |
| 5/27/2020 | 2.0 | 2.0 | 1 |

\*Number of isolated cases

### Model fitting

A pseudocode for Approximate Bayesian Computation Sequential Monte Carlo (ABC-SMC) is presented below adopting what was presented in the previous study (Minter and Retkute, 2019):

1. Set the number of generations *G* and the number of particles *N*
2. Set the tolerance schedule *ϵ*_1_ < *ϵ*_2_ < *ϵ*_3_ < ⋯ < *ϵ*_*G*_ and set the generation indicator *g* = 1
3. Set the particle indicator *i* = 1
4. If *g* = 1, sample *θ*^**^ from the prior distribution *p*(*θ*). If *g* > 1, sample *θ*^*^ from the previous generation {*θ*_*g*−1_} with weights {*w*_*g*−1_} and perturb the particle to obtain *θ*^**^∽*K*(*θ*|*θ*^*^)
5. If *p*(*θ*^**^) = 0, return to Step 4.
6. Generate *n* data sets 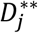 from the model using *θ*^**^ and calculate

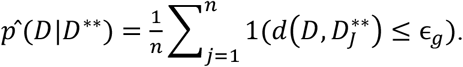
7. If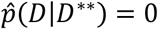 , return to Step 4
8. Set 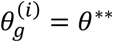 and calculate the corresponding weight of the accepted particle *i*

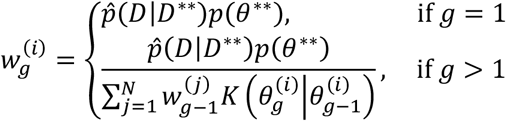
9. If *i* < *N*, increment *i* = *i* + 1 and go to step 4.
10. Normalize the weights so that 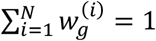
11. If *g* < *G*, set *g* = *g* + 1, go to step 3

*K*(*θ*|*θ*^*^) was assumed to follow a multivariate normal distribution that was truncated to give only positive values. Twenty generations (i.e., *G* = 20) were used with the following tolerance for Shincheonji 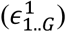 and non-Shincheonji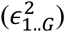 :

Initial tolerance values 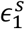 were set as

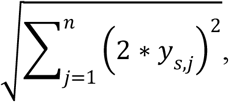

where *y*_*j*_ represent incidence of confirmed case on day *j* for Shincheonji (*S* = 1) and non-Shincheonji (*S* = 2), respectively. 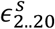 values were determined by setting the minimum values, 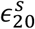 , as 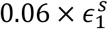 and dividing 20 equidistance pieces. Finally, final two value were manually adjusted to provide good fit between data and the model predictions through trial and error. Below are actual values used but were rounded for presentation.

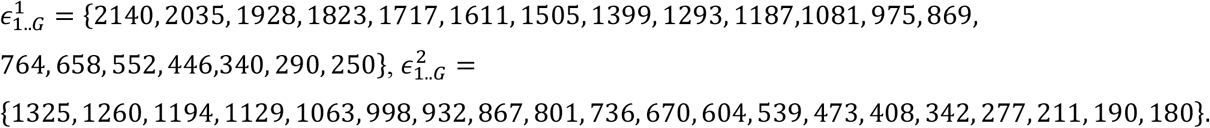

Prior distributions for the parameters *θ* = (*R*_0,1_, *R*_0,2_*I*_0_, *c*_12_, *d, R*^final^) were defined as uniform distribution as follows: *R*_0,1_ ∽ *U*(1, 20), *R*_0,2_ ∽ *U*(1, 20), *I*_0_ ∽ *U*(1, 20), *c*_12_ ∽ *U*(0.000001, 1), *d* ∽ *U*(1, 30), *R*^final^ ∽ *U*(1, 20).

**Figure S2.**
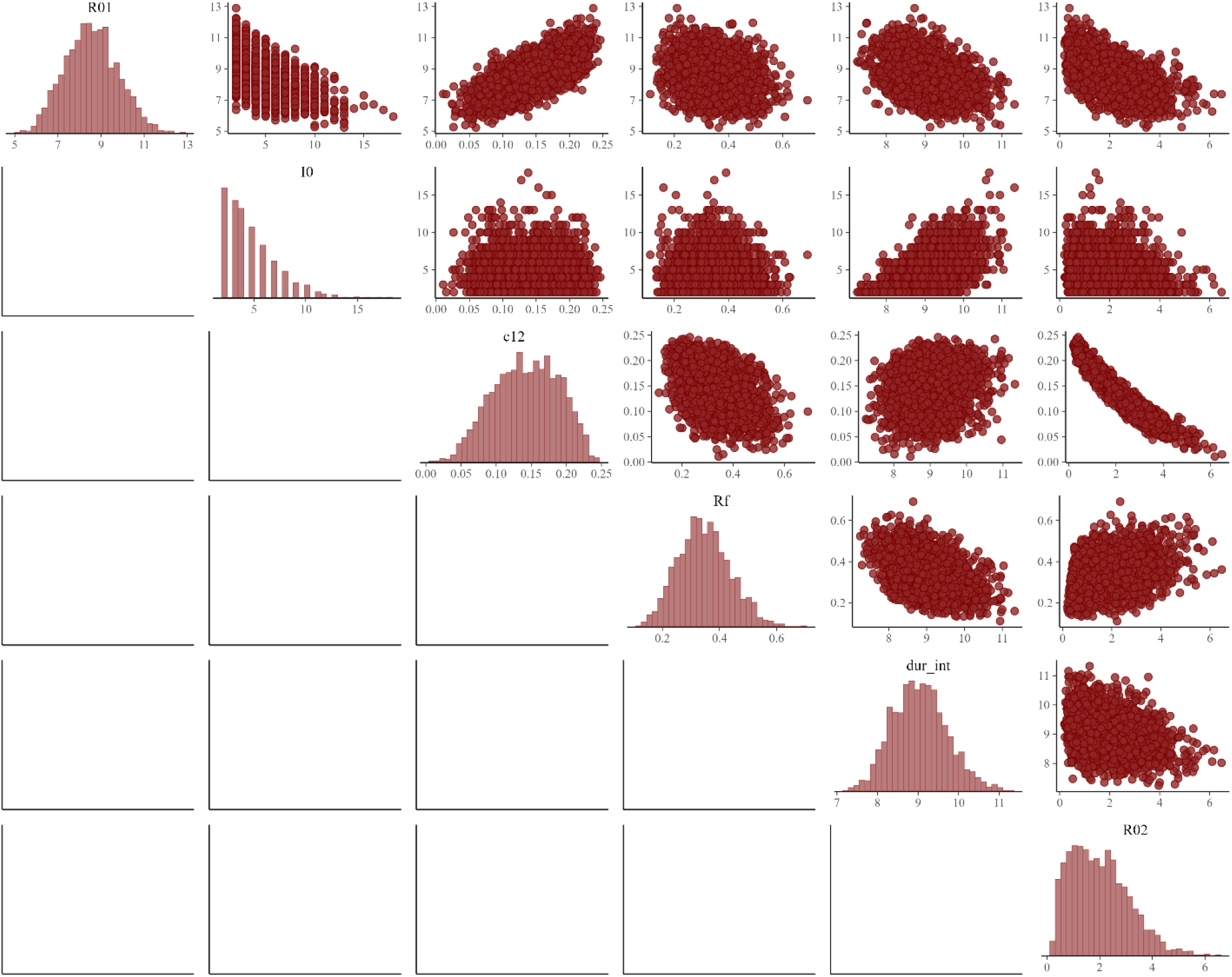
Posterior distribution of model parameters (*n*=2000). For each of 10 random seeds, 200 samples were generated.

### Growth rate *r* and basic reproduction number *R*_0_

For the differential equation-based *SIR* model, the initial (*i.e*., the entire population is susceptible) epidemic growth rate *r*\* can be given as *β* − *γ* (Ma, 2020), where *β* and *γ* represent transmission rate and recovery rate, respectively, as we defined in our model. Similarly, for a differential equation-based *SEIR* model *r*\* is given as below (Ma, 2020; Ma et al., 2014):

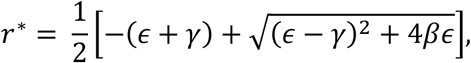

where *ϵ* represent the rate at which the exposed individuals become infectious (*i.e*., 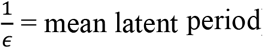) as we defined in the main text. The above equation gives *β* and therefore *R*_0_ for given *r*\*, *γ, ϵ*. Assuming *r*\* = *r*, which is the growth rate we calculated in the main text, we can see to what value of *R*_0_ the doubling times we calculated in the main text are translated and qualitatively see if *R*_0_ estimates from the current study are reasonable.

**Table S2.** Basic reproduction number, *R*_0_, calculated by assuming the empirical daily or weekly growth rate *r* is the same as the *r*\* calculated for the SEIR model.

| Date | $*R_0$ calculated from the<br>daily doubling time | | $R_0$ calculated from the<br>weekly rolling doubling time | |
| --- | --- | --- | --- | --- |
|  | Shincheonji | Non-Shincheonji | Shincheonji | Non-Shincheonji |
| 2020-02-18 | - | - | - | - |
| 2020-02-19 | 116.0 | - | - | - |
| 2020-02-20 | 33.0 | 51.6 | - | - |
| 2020-02-21 | 24.7 | 17.0 | - | - |
| 2020-02-22 | 17.8 | 4.1 | - | - |
| 2020-02-23 | 14.0 | 35.0 | - | - |
| 2020-02-24 | 9.5 | 2.1 | - | - |
| 2020-02-25 | 1.9 | 6.3 | 23.5 |  |
| 2020-02-26 | 3.8 | 15.4 | 12.9 | 15.1 |
| 2020-02-27 | 5.8 | 15.3 | 9.7 | 11.6 |
| 2020-02-28 | 6.6 | 1.5 | 7.7 | 9.0 |
| 2020-02-29 | 6.6 | 16.7 | 6.4 | 10.8 |
| 2020-03-01 | 6.1 | 1.1 | 5.5 | 6.8 |
| 2020-03-02 | 2.7 | 5.2 | 4.6 | 7.4 |
| 2020-03-03 | 1.9 | 5.7 | 4.6 | 7.3 |
| 2020-03-04 | 2.1 | 2.5 | 4.3 | 5.6 |
| 2020-03-05 | 2.4 | 1.8 | 3.8 | 4.0 |

**Figure S3.**
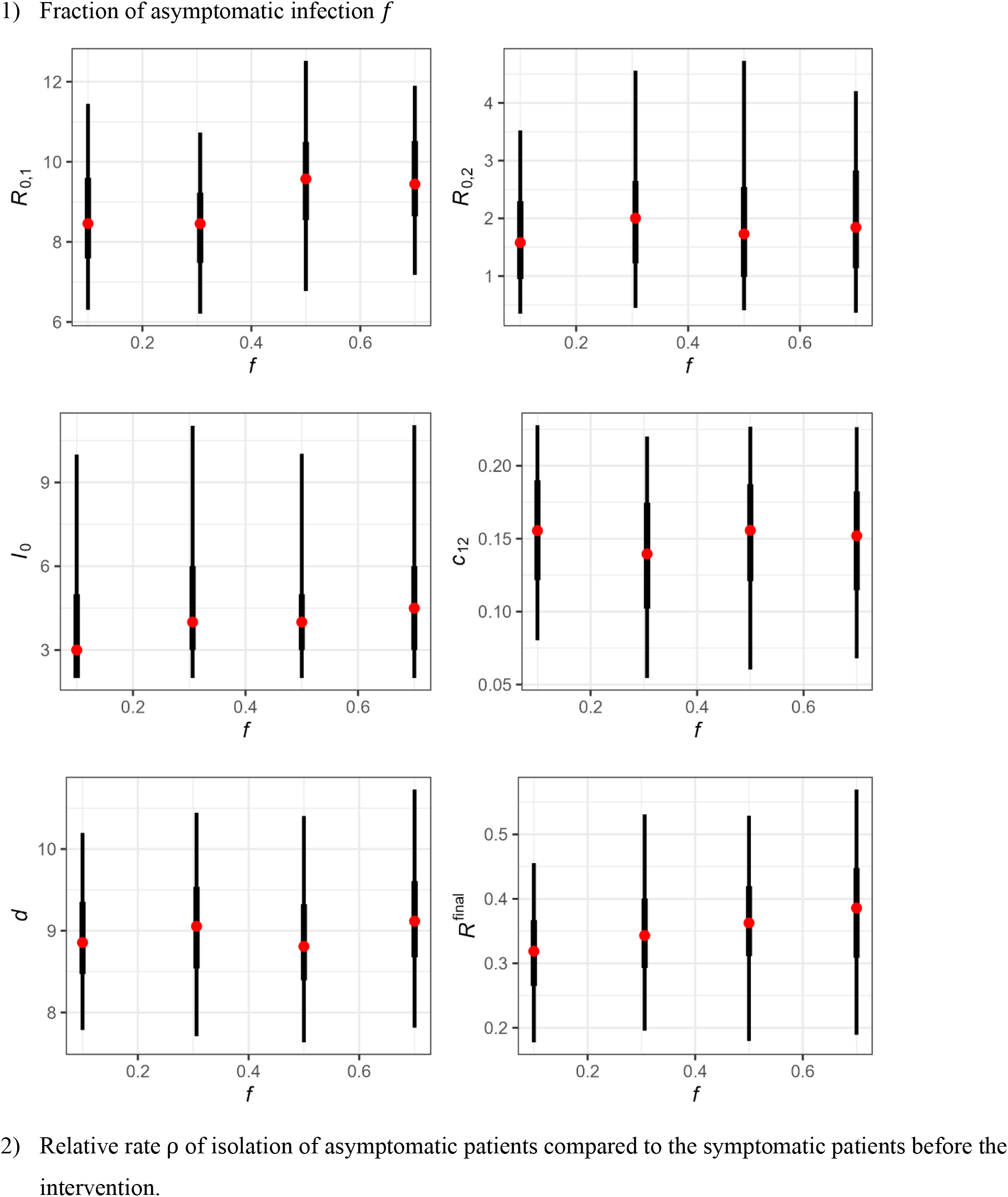
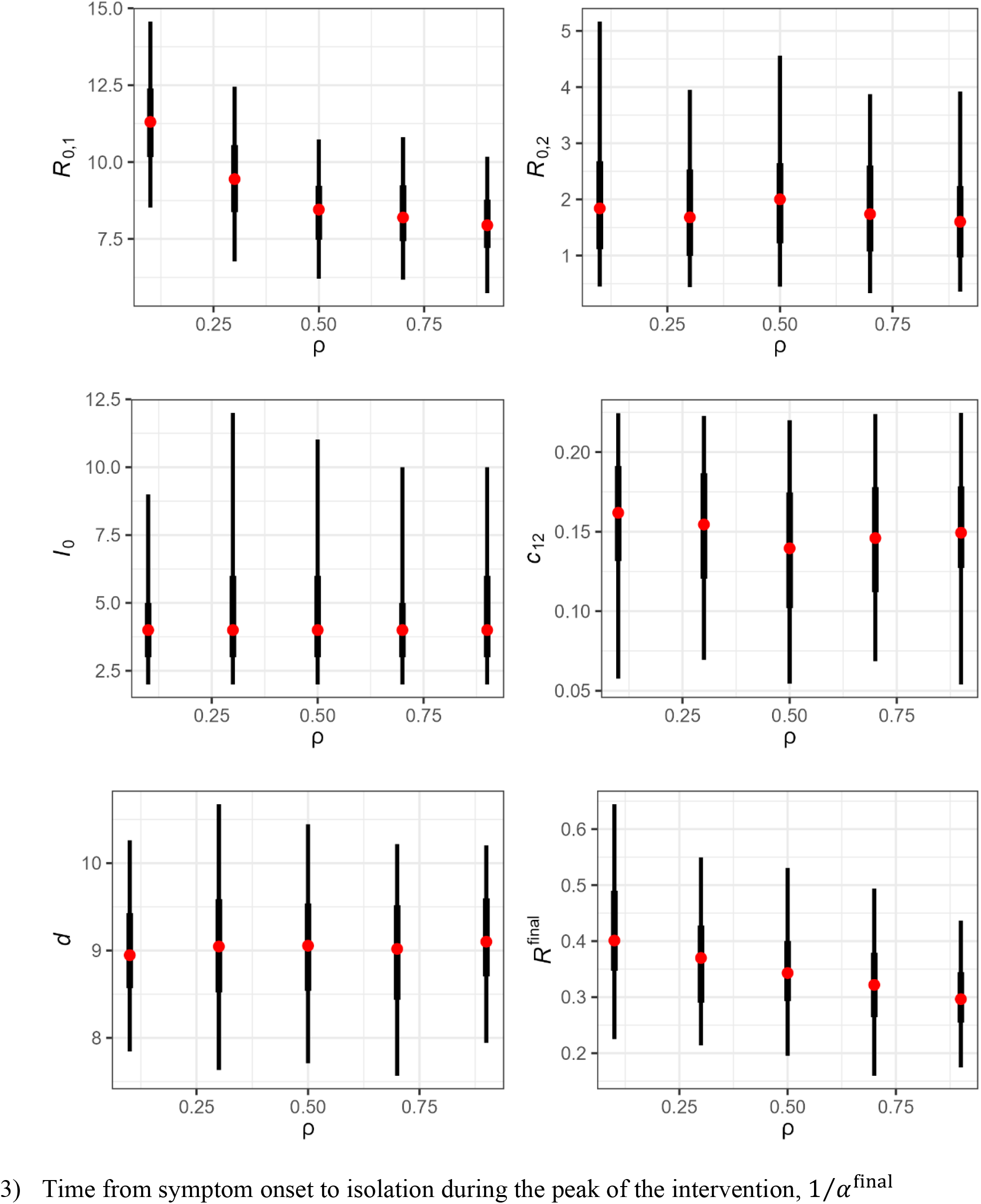
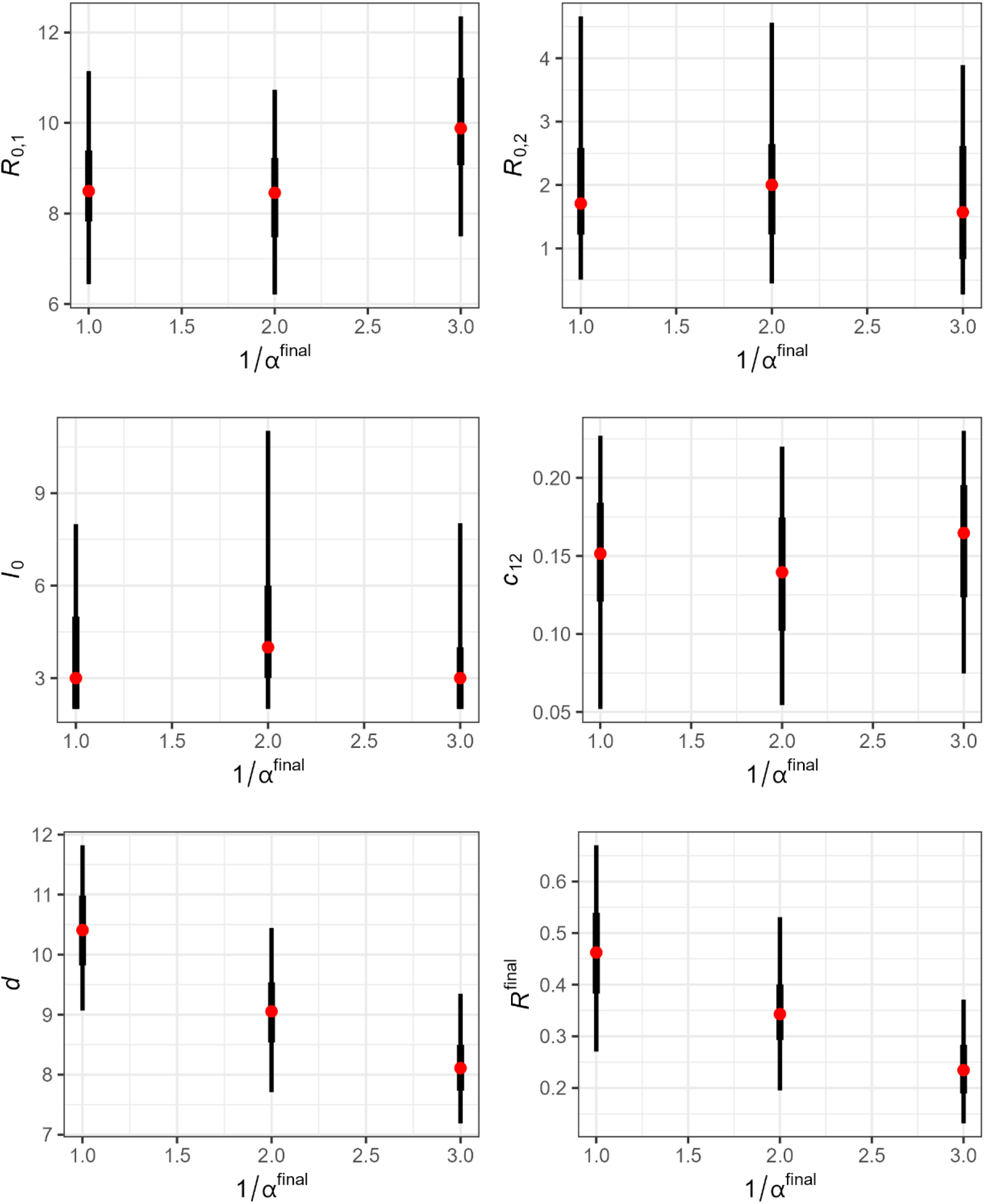
Sensitivity of our parameter estimates to simplifying assumption of three selected parameters

## Notes

### Competing Interest Statement

The authors have declared no competing interest.

### Funding Statement

This research was partly supported by Government-wide R&D Fund project for infectious
342 disease research (GFID), Republic of Korea (grant number: HG18C0088) and National Institute for
343 Mathematical Sciences (NIMS) grant funded by the Korean Government (NIMS-B21910000).

### Author Declarations

This study uses publicly available data and would not require IRB approval.

### Summary of Updates

Author affiliations updated; Supplemental files updated.

